# The impact of early public health interventions on SARS-CoV-2 transmission and evolution

**DOI:** 10.1101/2020.11.18.20233767

**Authors:** Sebastian Duchene, Leo Featherstone, Birgitte Freiesleben de Blasio, Edward C. Holmes, Jon Bohlin, John H.-O. Pettersson

**Affiliations:** Department of Microbiology and Immunology, The University of Melbourne at The Peter Doherty Institute for Infection and Immunity, Melbourne, Victoria, Australia; Department of Methods Development and Analytics, Division of Infectious Disease Control and Environmental Health, Norwegian Institute of Public Health, Oslo, Norway; Department of Biostatistics, Oslo Centre for Biostatistics and Epidemiology, Institute of Basic Medical Sciences, University of Oslo, Norway; Marie Bashir Institute for Infectious Diseases and Biosecurity, School of Life and Environmental Sciences and School of Medical Sciences, the University of Sydney, Sydney, New South Wales 2006, Australia; Zoonosis Science Center, Department of Medical Biochemistry and Microbiology, University of Uppsala, SE-751 23 Uppsala, Sweden

**Keywords:** SARS-CoV-2, Nordic, transmission chain modelling, virus exportation

## Abstract

**Background:** Many countries have attempted to mitigate and control COVID-19 through the implementation of non-pharmaceutical interventions, particularly with the aim of reducing population movement and contact. However, it remains unclear how the different control strategies impacted the local phylodynamics of the causative SARS-CoV-2 virus.

**Aim:** To assess the duration of chains of virus transmission within individual countries and the extent to which countries export viruses to their geographic neighbours.

**Methods:** To address core questions in genomic epidemiology and public health we analysed complete SARS-CoV-2 genomes to infer the relative frequencies of virus importation and exportation, as well as virus transmission dynamics, within countries of northern Europe. To this end, we examined virus evolution and phylodynamics in Denmark, Finland, Iceland, Norway and Sweden during the first year of the pandemic.

**Results:** The Nordic countries differed markedly in the invasiveness of control strategies implemented. In particular, Sweden did not initially employ any strict population movement limitations and experienced markedly different transmission chain dynamics, which were more numerous and tended to have more cases, a set of features that increased with time during the first eight months of 2020.

**Conclusion:** Together with Denmark, Sweden was also characterised as a net exporter of SARS-CoV-2. Hence, Sweden effectively constituted an epidemiological and evolutionary ‘refugia’ that enabled the virus to maintain active transmission and spread to other geographic localities. In sum, our analysis reveals the utility of genomic surveillance where active transmission chain monitoring is a key metric.

## Introduction

Since its initial description in December 2019, SARS-CoV-2, the causative agent of COVID19 [1, 2], has rapidly led to a major global health crisis. The pandemic has caused more than 150 million infections and over 3.3 million deaths worldwide and continues to accelerate, imposing a significant impact on health care systems, societies and the global economy. Countries are continuously struggling with how to effectively counteract the pandemic, balancing the protection of health with social and economic considerations.During the first year of the pandemic, in the absence of therapeutics and available therapies and vaccines, efforts centred on so-called ‘non-pharmaceutical strategies’, particularly initial short-term large-scale restrictions to population movement (e.g. ‘lock-downs’), increased testing and various levels of social distancing. Analysis of the local epidemiological consequences of different control strategies provides key information on the most effective approaches to reduce the rate of virus transmission within communities.

The Nordic countries, defined here as Denmark, Finland, Norway, Iceland and Sweden, provide a powerful example of geographically, politically and socially related countries that differ markedly in how control strategies for COVID-19 were implemented during the first six months of the pandemic. In particular, Sweden initially took a less restrictive approach in which no general population movement restrictions were enforced, schools for children below the age of 16 years old remained open, no mandatory quarantine was imposed for infected households, and businesses continued almost as usual [3]. In contrast, Norway and Denmark enforced a more invasive population movement restriction that included enforced home office for workers in the public sector, home-bound schooling, targeted private sector close downs, as well as closed international borders for non-residents. Iceland, a relatively small homogenous island population (of ∼350,000) never initiated a population movement restriction as Norway and Denmark, but rather focused on large-scale testing and contact tracing to limit virus spread within the community. In relation to population size, Sweden has had a higher number of COVID19-related cases and deaths than all other Nordic countries [4, 5], with a total of around 9,758/137 cases/deaths per 100,000 people, compared to around 4,465/43 in Denmark, 2,169/14 in Norway, 1,608/17 in Finland, and 1,790/8 in Iceland, as of 12^th^ of May 2021 [6], with most cases between April and July 2020 and November 2020 to March 2021.

Although the relative ‘success’ of COVID-19 control measures are normally gauged in the number of cases and deaths at the country level, it is also the case that intervention and mitigation strategies may lead to marked differences in transmission dynamics among populations, which may in turn impact the evolution of the virus. Using a comparative analysis of genome sequence data we addressed whether the different approaches to COVID-19 control employed by the Nordic countries resulted in differences in virus transmission dynamics and the relative frequencies of virus importation/exportation (i.e. virus “phylodynamics”) during the first year of the pandemic following introduction of SARS-CoV-2 in the Nordic region. Accordingly, we compiled an extensive data set of SARS-CoV-2 virus genomes and performed a phylo-epidemiological study to identify any differences in transmission chain dynamics between these countries.

## Materials and Methods

### Data set construction

We downloaded all SARS-CoV-2 genomes that were complete and had high sequencing coverage available the 22^nd^ March 2021 at the GISAID platform (www.gisaid.org), for which no ethical approval was needed. We selected all those from the five Nordic countries as follows: 50,126 from Denmark, 2,374 from Finland, 4,167 from Iceland, 3,388 from Norway, and 7,863 from Swede). To obtain a representative subset of the global diversity we also selected 3,437 genomes from the latest NextStrain global build in 22^nd^ March [7].

Sequencing intensity is markedly different between countries, with Iceland sequencing around 68% of positive reported cases whereas in Sweden the corresponding number is about 1.1%. We attempted to control for such sampling bias by subsampling genomes from the Nordic countries relative to that with the lowest sampling intensity, meaning that all countries were sampled a rate of 1.1 genomes per hundred cases. For this purpose, we divided the number of genomes from each country by the cumulative number of cases reported in each country by mid-March 2021, as recorded in ourworldindata.org [8]. We repeated this procedure 10 times, and in each obtained a sequence alignment with all the data from NextStrain and with the number of genomes from the Nordic countries proportional to the prevalence of the virus (Table 1). The complete data set consisted of 71,355 genomes, with 67,918 from the Nordic countries. Our 10 alignments with equal sequencing intensity for Nordic countries had between 15,616 and 15,297 sequences (i.e. between 12,179 and 11,860 Nordic genomes, plus the 3,467 from the NextStrain build). The GISAID accession numbers of all sequences included in the final sequence alignment are available as Supplementary Table S1.

**Table 1.**
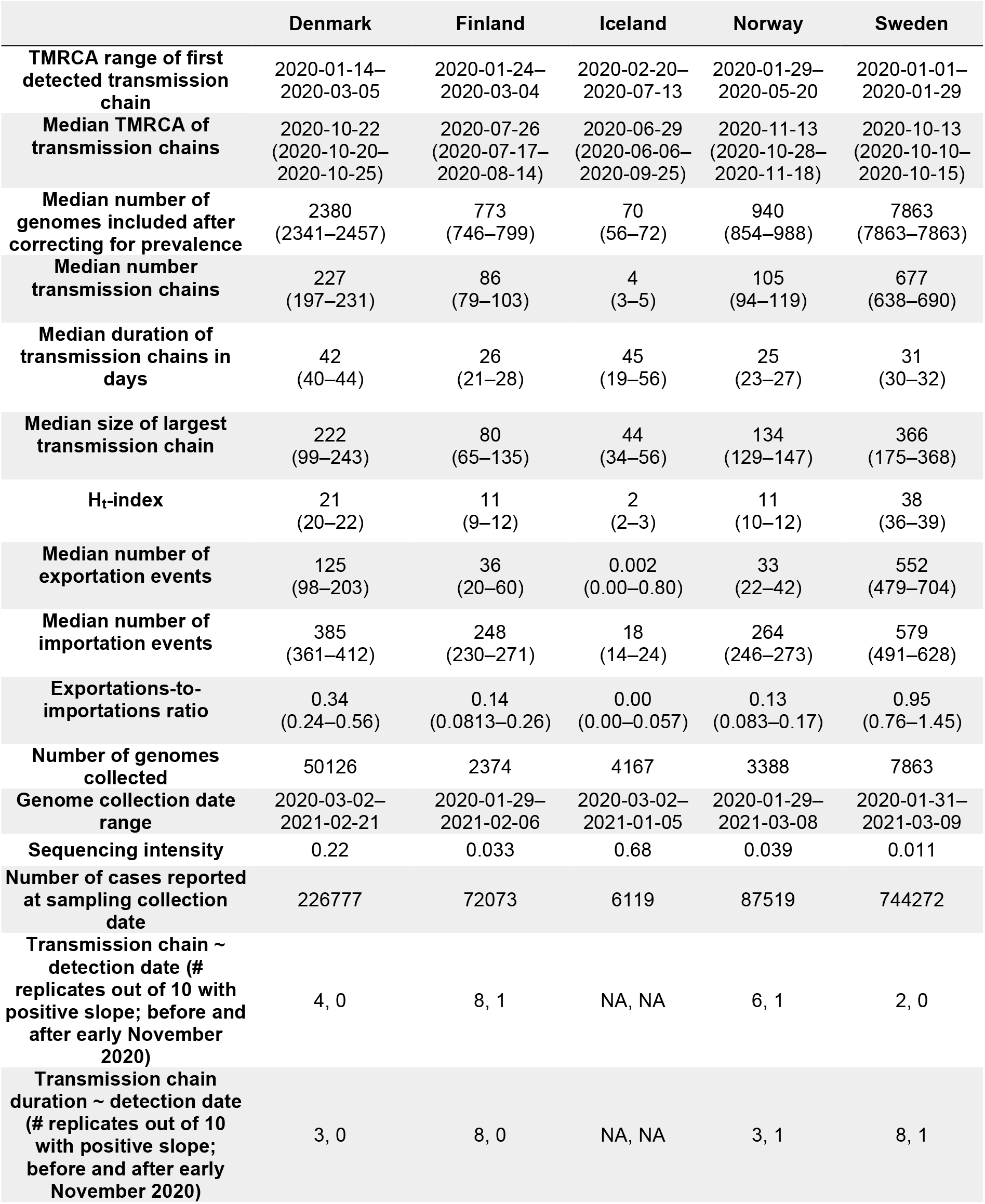
Summary statistics computed for SARS-CoV-2 data from each Nordic country. The median and range (over 10 replicates) is shown for relevant statistics, with the exception of the time to the most recent common ancestor (TMRCA; the putative time of emergence) of the first transmission chain, for which only the range is reported because the average disproportionally affects countries with low prevalence. For regression statistics, the symbol ∼ denotes “function of”, and in these cases we report the number of replicates with positive regression slopes before and after 31^st^ August 2020.

### Estimating a time-scaled phylogenetic tree

We estimated maximum likelihood phylogenetic trees using IQ-TREE v2.0.6 [9] employing the GTR+ Γ model of nucleotide substitution [10]. We fit a strict molecular clock to the data using LSDv0.3 [11]. We calibrated the molecular clock by specifying the sequence sampling times and fixing the evolutionary rate to 1×10^−3^ subs/site/year as estimated previously [12]. We chose this approach because testing for temporal signal and fitting a sophisticated molecular clock model is unfeasible for a data set of this size. For a small number of sequences (113, ∼0.14% of the data set) only the month and year were available, such that we assigned them the 15^th^ of the corresponding month.

### Detection of virus transmission chains

We defined transmission chains as monophyletic groups of genomes (at least 2) all collected from one of the Nordic countries, which is analogous to transmission lineages as defined previously [13]. We computed key statistics from each transmission chain, specifically: the duration (i.e. the length of time in days from the first to the last collected genome), the size (i.e. the number of genomes), and the time of origin. For each country we also calculated an Ht-index of transmission chains, where Ht is the number of transmission chains with at least Ht cases (analogous to the H-index citation metric). Larger values reflect increase in the number and size of lineages. For example, of Sweden’s lineages, 38 had at least 38 cases (Table 1). Because we subsampled the data 10 times, for these statistics we compute the range of values, as a crude measure of uncertainty due to sampling.

We fit linear regressions for transmission chain size against as a function of the detection date (the date of collection of the first genome within a transmission chain), such that positive trends would suggest increased community spread over time (i.e. larger transmission chains over time). We also considered the duration of transmission chains as a function of detection date, where a positive trend means that transmission chains detected later in the pandemic tended to last longer. Because the number of cases had substantially decreased around August 2020 (Supplementary Figure S1) and because around this time most stringent public health interventions were in place (Supplementary Figure S2–S5), we fit our regressions across two periods, before and after 31^st^ August 2020. Importantly, this time also preceded the emergence of lineage B.1.1.7 and its introduction to the Nordic countries by a few months (Supplementary Figure S6). This so-called ‘variant of concern’ has been shown to have increased transmissibility relative to circulating diversity [14]. We only report trends as residual distributions were strongly skewed..

### Inferring virus migration dynamics between Nordic countries

To infer the frequency of importation or exportation events of SARS-CoV-2 between the Nordic countries and from the rest of the world, we employed a Bayesian stochastic mapping approach, also known as discrete phylogeography, as implemented in BEAST v1.10.4 [15–17] using guidelines from Dellicour et al. for very large genomic data sets [18]. We fixed the time-tree described above and assigned a geographic location for each tip, which could be either of the five Nordic countries or “other” (for those collected in other countries). This method is broadly similar to that used to infer geographic movement of the virus in Belgium [12]. We repeated this procedure for the 10 subsampled replicate trees and report median posterior values and ranges across replicates. Stochastic mapping generates a posterior distribution of ‘type-changes’ between locations along the branches.

We ran a Markov chain Monte Carlo of length 5×10^7^ steps, recording every 5000^th^ step. Sufficient sampling from the posterior was assessed by verifying that the effective sample size of all parameters were above 200 as estimated in Tracer v.1.7 [19]. We inferred the posterior number of migration events between the six possible states (Markov jumps) and the amount of time spent at each state, known as Markov rewards.

### Assessing public health measures and governmental stringency

To provide an overview of public health measures taken and how they were enforced between February 2020 through March 2021 per Nordic country, we plotted data from the Oxford COVID-19 Government Response Tracker [20] which included: testing policy, contact tracing, public information campaigns, international travel control, workplace closure, school closure, cancelation of public transportation, stay at home requirements, facemask usage, gathering restrictions, cancelation of public events, restrictions on movement, and government stringency response index (Supplementary figures S2–S5).

## Results

The estimated time of emergence of sampled transmission chains provides information about the onset of community transmission, although such an association is sensitive to sampling bias, particularly the time-scale of country-specific sequencing efforts. Our phylogenetic analyses revealed that SARS-CoV-2 was imported to all Nordic countries between January to late February 2020, with detectable community transmission from early February in Sweden and from early March in the other Nordic countries as inferred by the presence of transmission chains (Figure 1). Sustained community transmission continued for all Nordic countries beyond April, with the exception of Iceland which was characterised by a relatively short one-month period of community transmission, and Norway, which had a low number of cases combined with a low sequencing intensity of 0.039 (the second lowest in our study after Sweden; Table 1).

**Figure 1.**
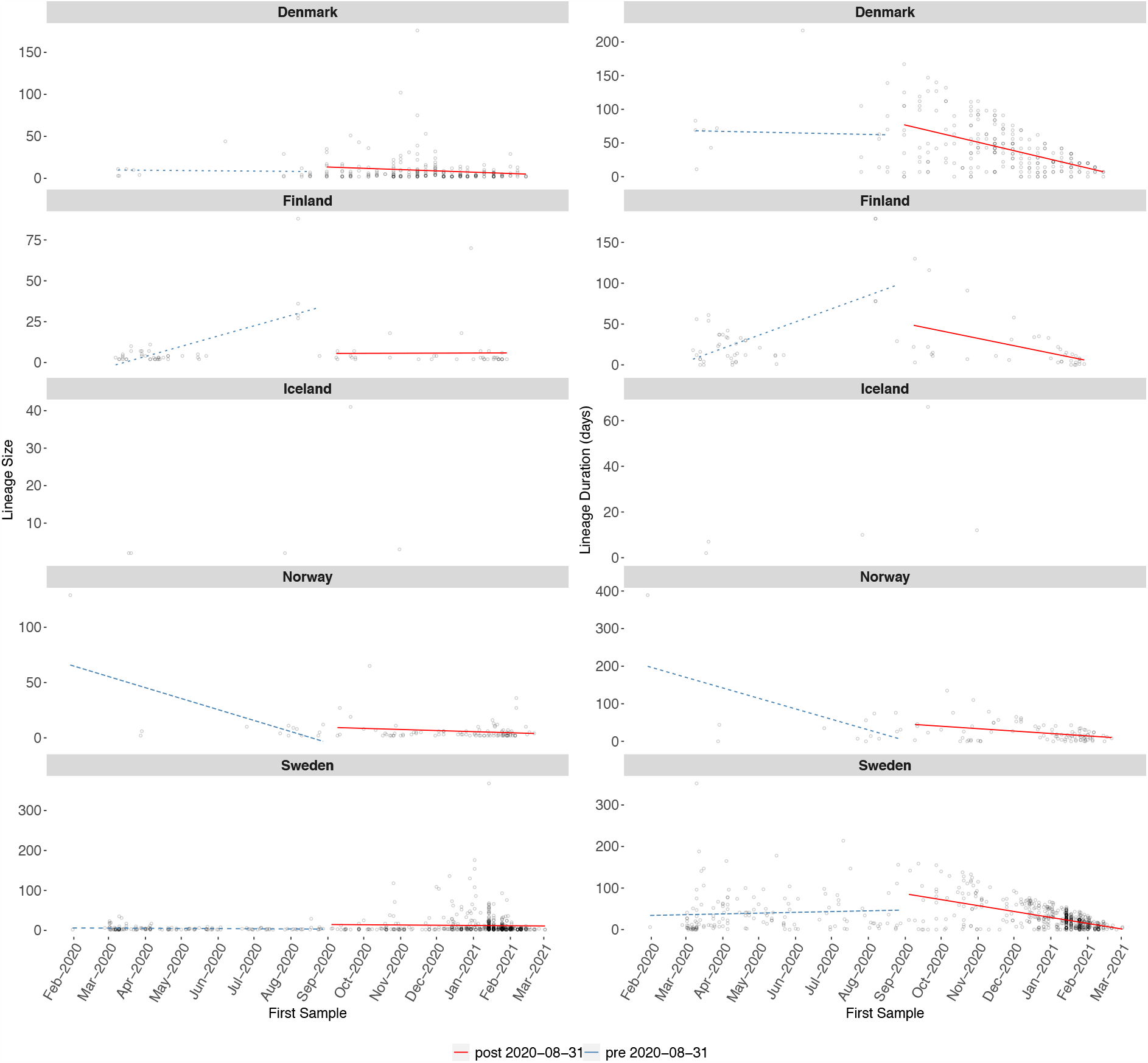
Transmission chain summaries. Open circles mark transmission chain size (left) and chain duration (right) against the time of the first sample from each chain for each country. Trend lines are fitted here to one of ten replicates with dotted lines corresponding to trends that were not repeated in at least 9 of 10 replicates. Trends are omitted for Iceland due to low sample size. Trends are split on either side of 31^st^ August 2020.

The number of transmission chains observed was around 227 (range: 197–231) for Denmark, 86 (range: 79–103) for Finland, in Iceland (45, range: 19–56), and 105 (range: 94– 119) for Norway. Notably, the highest number of transmission chains - 677 (range: 638–690) - was observed in Sweden. The median duration of such transmission chains (number of days from detection to last sampled case) was higher in Iceland (45, range: 19–56) and Denmark (42, range: 40–44) than other localities: 31 (range: 30–32) for Sweden, 26 (range:21–28) for Finland, and 25 (range: 23–27) for Norway (Table 1).

The largest transmission chain was found in Sweden, with 366 cases (range: 175–368), followed by Denmark with 228 (range: 99–243), Norway with 134 (range: 129–147), Finland with 80 (range: 65–135), and Iceland with 44 (range: 34–56). In addition to having the largest transmission chain, Sweden also had the highest Ht-index (Table 1). The Ht-index for Sweden was 38 (range: 36–39), 21 for Denmark (range: 20–22), 11 for Finland (range: 9– 12), 2 for Iceland (range: 2–3), and 11 for Norway (range: 10–12), suggesting that Sweden had the least-interrupted community transmission.

There was inconclusive evidence of an increase in transmission chain size in the first period (prior to the 31^st^ August 2020), with between 2 and 8 of 10 subsampling replicates displaying a positive regression slope. Due to low sample size and number of data points in the second period we did not perform a trend analysis for Iceland. After 31^st^ August 2020 most replicates (9 or more) exhibited a negative trend for all countries, indicating a decrease in transmission chain size over time. Similarly, we did not find consistent trends for the duration of transmission chains in the first period, whereas in the second period they appeared to decline over time, with at most one replicate displaying a positive slope in Denmark, Finland, Norway and Sweden (Table 1; Figure 1). Overall, these results are consistent with stringent public health interventions in the second period examined here that may have limited the spread of existing transmission chains. Thus, the increased number of cases observed in the second half of 2020 was most likely driven by importations that led to limited or ceased transmission, rather than an increasing size and duration of transmission chains. This is consistent with the majority of importation events leading to transmission chains or singletons coming in the second half of 2020 for most countries (Supplementary Figure S7).

The total number of exportation events was larger for Sweden (552, range: 479–704) than for Denmark (125, range: 98–203). Most exportation events from Sweden were into Finland, whereas those from Denmark were into non-Nordic countries, followed by Sweden and Iceland (Figure 2, Panel B). An inspection of the amount of time in the tree occupied by each country, known as the Markov rewards, revealed that Sweden occupied the largest portion, consistent with it being the major exporter. In contrast, Norway, had very low Markov rewards, despite having more genomes than Iceland and Finland, both with higher Markov rewards (Supplementary Figure S8).

**Figure 2.**
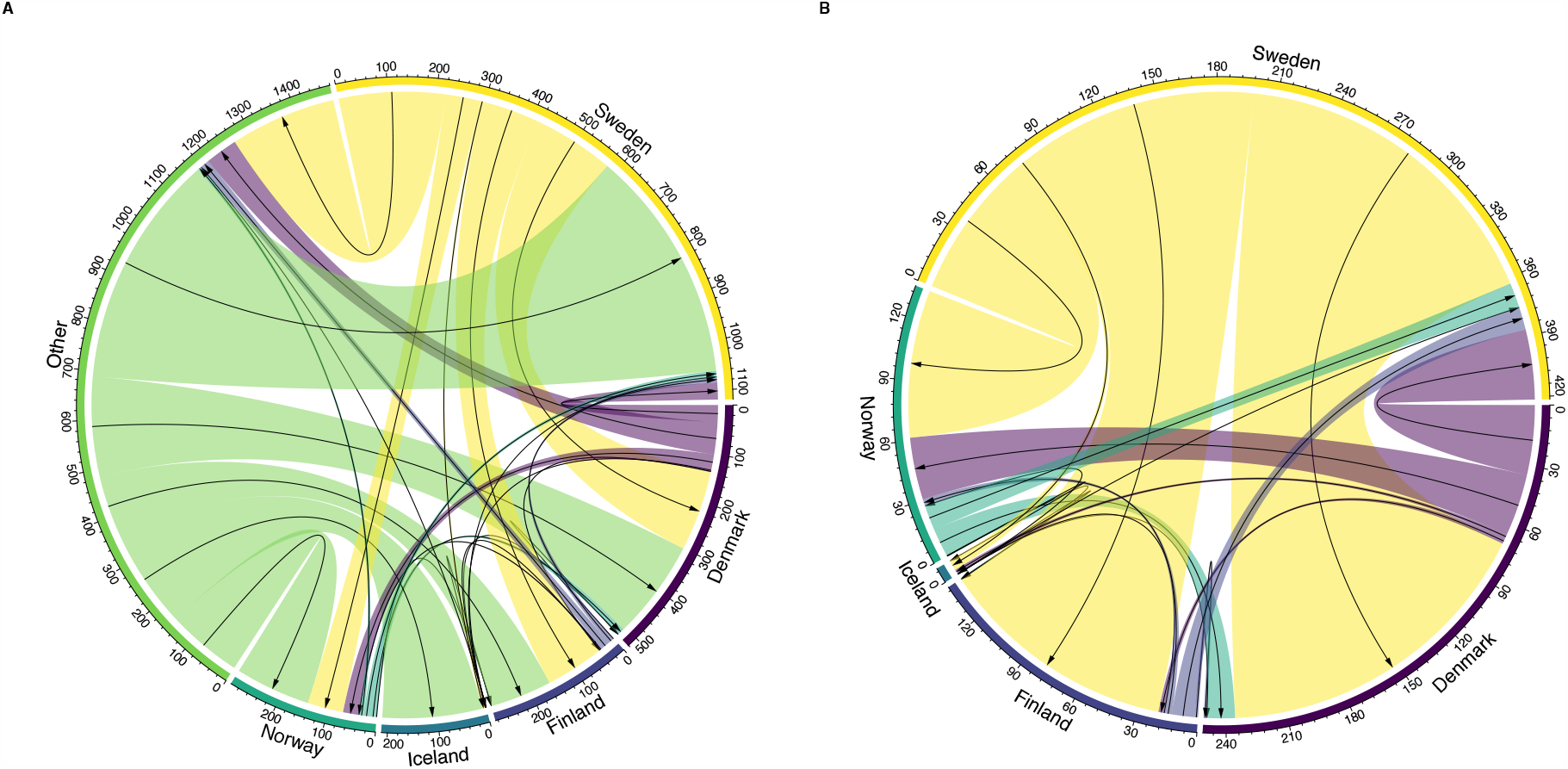
Circular migration diagram of migration events between the five Nordic countries and other locations. Panel A shows migration events with ‘Other’ countries included and Panel B shows migration between Nordic countries only. The size of the coloured links denotes the posterior median number of inferred migration events from the Bayesian phylogeographic analysis across 10 replicates. The black arrows represent the migration direction.

## Discussion

Until March 2021, Sweden experienced a greater number of COVID-19-related cases and deaths in relation to population size than all other Nordic countries [6]. Our genomic analysis shows that whilst SARS-CoV-2 was imported to all Nordic countries during the same time-period in early 2020, Sweden experienced more transmission chains which also tended to be larger. Importantly, these trends emerged despite Sweden having the lowest virus genome sampling proportion, suggesting that community transmission was well-established during both waves relative to the other Nordic countries. Reassuringly, all Nordic countries displayed a decrease in the size of transmission chains and their duration in the second time period examined here.

Denmark and Sweden were characterised by having acted as exporters of SARS-CoV-2 to their Nordic neighbours. Conversely, Iceland, Norway and in particular Finland showed lower levels of virus exportation. At the same time, our analyses indicate that Finland was the main receiver of the exportations from Sweden, suggesting a near unidirectional mode of exportation between the two countries.

During the first six months of the pandemic Sweden never enforced strict community mobility restrictions. Instead, social distancing, limiting sizes of social events and moving to distance education for students above 16 years were employed as the main mitigation efforts. Conversely, Denmark, Finland and Norway enforced a relatively strict population movement restrictions of their communities, closure of borders and government run facilities [4]. These mitigation efforts were taken during both the first and second wave (Supplementary table S2, Supplementary figures S2–S5). During the second wave, which started in early September 2020, Sweden recommended limiting the number of people that meet within social circles. In early 2021, Sweden also recommended the use of protective face-masks in public spaces. In Norway, locally-based contact tracing, testing, isolation, and quarantining, supported by a national contact tracing team, has been the primary strategy since the summer of 2020. In contrast, the danish strategy focuses on extensive testing, free-of-charge with readily accessible testing units (Supplementary figures S2). Both countries enforce local contact-reducing measures during periods of outbreaks. Iceland, in a similar manner to South Korea, utilised large-scale testing and contact tracing combined with social distancing and voluntary home-based quarantine without the need to regulate population movement [21] (see also Supplementary table S2). Thus, while regulating population movement appears to provide an efficient mode to reduce community transmission [22, 23], other mitigation efforts are also viable options under certain circumstances. Norway experienced relatively low numbers of transmission chains during the first six months of the pandemic, with the bulk of the transmission chains sampled here emerging in mid-April and towards the end of July. Although this might in part reflect a delayed start to genome sequencing, which have increased significantly in all countries since then, and such results should be interpreted with caution (Figure 1). Indeed, we note that the sequencing intensity of Norwegian cases is lower than some of the other Nordic countries with similar case burdens (Table 1). Thus, sequencing proportion bias per country needs to be taken into account when considering our results. To reduce effect of sequencing bias, we attempted to subsample our data to obtain even sequencing proportions although this does not account for other sources of bias, such as if close contacts of a positive case are sequenced or not. When sample size per country increases with time, the effect of such potential sampling biases are expected to decrease.

To what extent then did Sweden’s mitigation strategy affect the epidemiological situation internally and across Nordic region as a whole? Sweden received substantial attention due to the number of deaths reported. Sweden has been shown to have had one of the longest durations of excess deaths and highest case numbers between February and May 2020 [5] and it has been suggested that a greater level of self-isolation likely would have reduced the number of deaths in Sweden [24].

The effect of closing/opening schools are also likely to have contributed to the transmission dynamics [25, 26]. In the case of the Nordic countries, it can be argued that by closing pre-, primary, secondary schools and higher education for several weeks during the spring of 2020 (Supplementary table S2, Supplementary figure S3), Denmark and Norway also reduced contact and virus transmission in the adult population to a higher extent than Sweden where schools remained open during the same period. However, although Sweden exhibited a relatively high number of cases compared to the other Nordic countries, and our results are consistent with a relatively high community transmission for Sweden during the first twelve months, such an increase in community transmission does not necessarily imply a causal relationship with excess deaths. Whilst we employ a robust genomic-based modelling approach to study transmission chain changes, equating or directly relating such changes with the number of deaths may lead to incorrect conclusions as we have not taken into account any patient-related epidemiological parameters such as gender or age. Furthermore, it has been argued that the higher number of deaths in Sweden may also have been influenced by how intensive care unit admission criteria were applied, particularly for the elderly population in care homes, during the early phase of the pandemic [24]. Thus, official mitigation strategies and efforts alone are unlikely to explain epidemiological differences between countries, which were also impacted by the compliance and practical enforcement from the general public, businesses, and health care providers.

A critical issue, although one that has received little attention, is that if transmission chains are allowed to remain active, they also provide increased opportunity for the virus to evolve and adapt to local populations, potentially acquiring mutations that in some way enhance virus fitness. Indeed, several so-called ‘variants of concern’ have independently emerged globally with mutations that are associated with epidemiologically important properties. For example, the B.1.1.7 variant has acquired mutations that has significantly increased the transmission potential of the virus [14] and the B.1.351 variant has shown increased resistance against vaccines [27]. Clearly, mutations routinely appear in the SARS-CoV-2 genome, including viruses sampled from Sweden [28], and it is important to continuously evaluate their functional relevance. In addition to providing increased evolutionary potential, sustained transmission chains also provide epidemiological ‘refugia’ for the virus to be transmitted to other localities. This is in line with our results showing that Sweden had a greater frequency of exportation events among neighbouring Nordic countries. As a consequence, interrupting and ultimately stopping transmission chains is not only important to minimise virus spread within populations, but also to reduce the chances for the virus to accumulate beneficial mutations. Fortunately, COVID-19 vaccination has started in all Nordic countries around the New Year of 2020/2021, which will reduce transmission chains and protect vulnerable patient groups. However, genome sequencing will continue to play an important role to monitor for potential vaccine escape mutants.

Overall, our study highlights the utility of continuous genomic surveillance and retrospective studies to compare and understand differences in pandemic responses with respect to transmission dynamics. In particular, the data presented suggests that transmission chain monitoring may prove to be a useful metric in comparing outbreak mitigation outcome.

## Data Availability

Accession numbers to the sequence data used in the study (full data available at the GISAID platform) are available in supplementary table 2.

## Acknowledgements

We thank all people who have contributed SARS-CoV-2 genome data to the GISAID platform. Details on specific contributions are available via www.gisaid.org/. JHOP is funded by the Swedish research council FORMAS (grant no: 2015-710) and VR (grant no: 2020-02593). ECH is funded by an ARC Australian Laureate Fellowship (FL170100022). SD and LF are funded by an ARC Discovery Early Career Award awarded to SD (DE190100805).

## Supplementary materials for this manuscript including the following

**Supplementary Figure S1.**
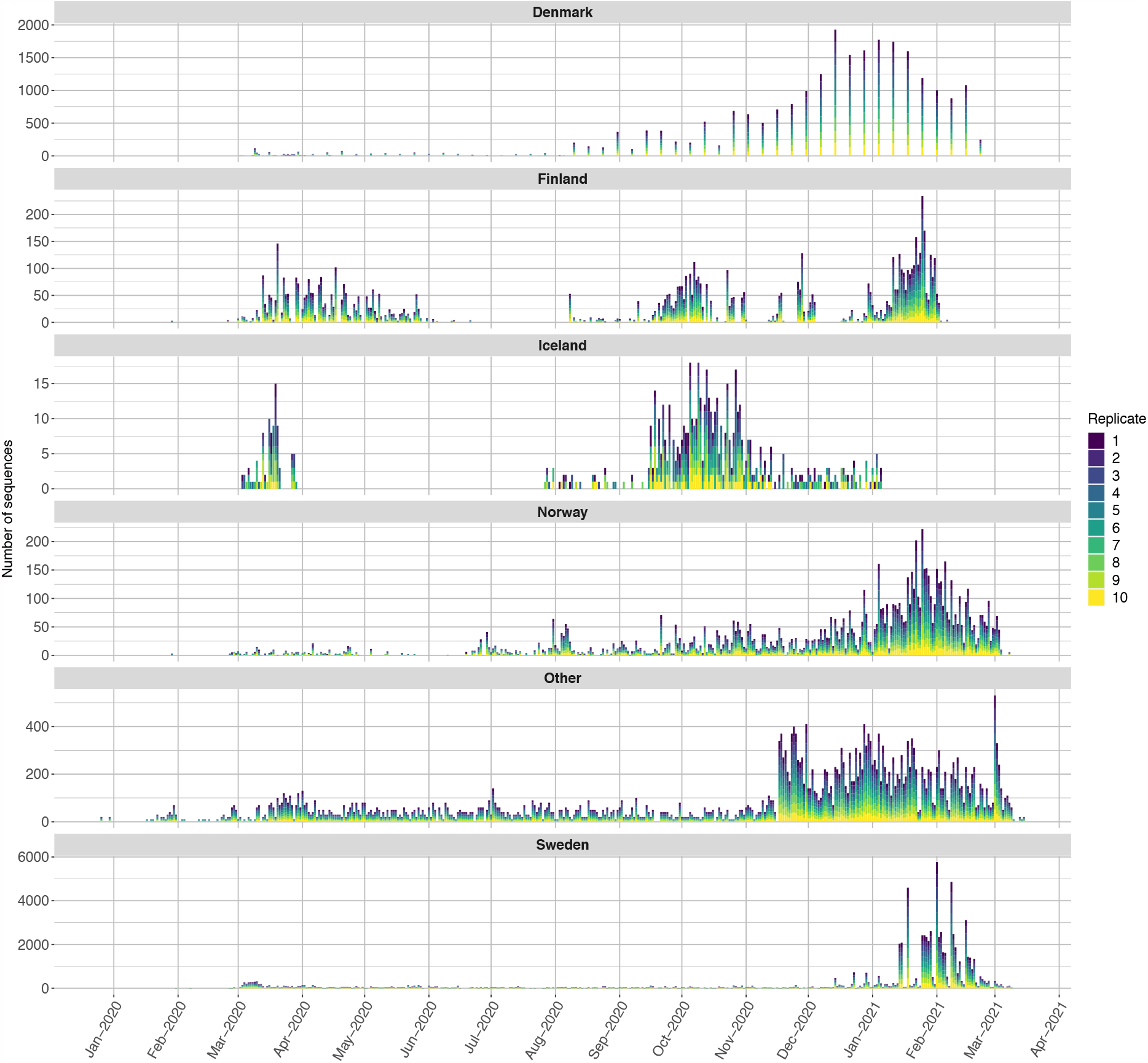
Number of SARS-CoV-2 genomes sampled with time for all Nordic countries and global samples (i.e. ‘other’) included in the final alignment.

**Supplementary Figure S2.**
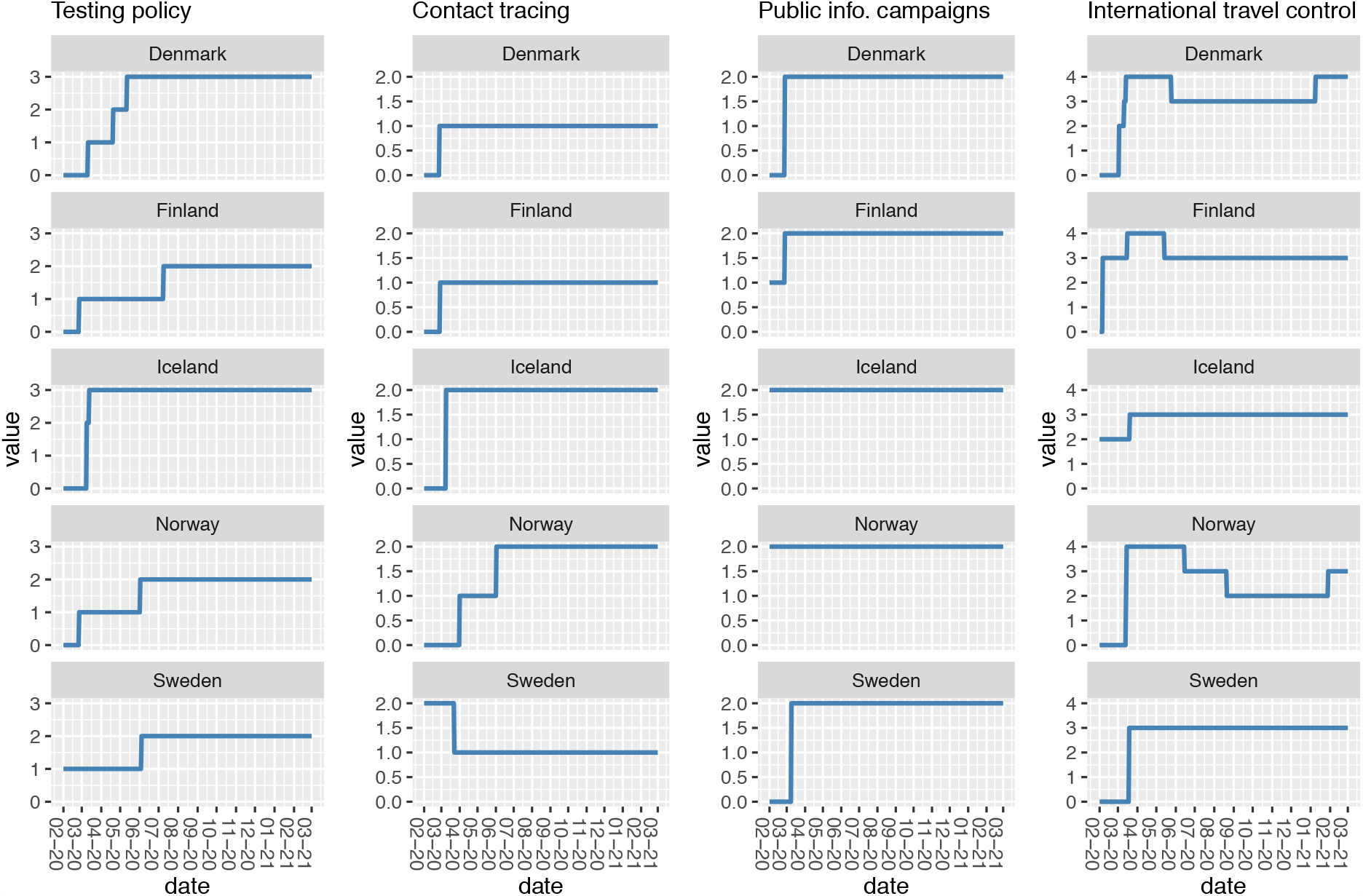
Information on indicators of government response between February 2020 through February 2021 by country regarding (i) testing policy, (ii) contact tracing, (iii) public information campaigns, and (iv) international travel control. All indicators are represented using ordinal scales, showing the highest value in the countries at each time point. If local measures are used, these values may apply to a limited geographic area. Testing policy: 0: no; 1: symptomatic and certain criteria; 2: anyone with symptoms; 3: open public testing; Contact tracing: 0: no; 1: limited; 2: comprehensive; Public information campaigns: 0: no; 1: public officials cautions; 2: coordinated campaigns; International travel control (foreign travellers exclusively): 0: no; 1: screening arrival; 2: quarantine arrival; 3 ban arrivals some regions; 4: total border closure.

**Supplementary Figure S3.**
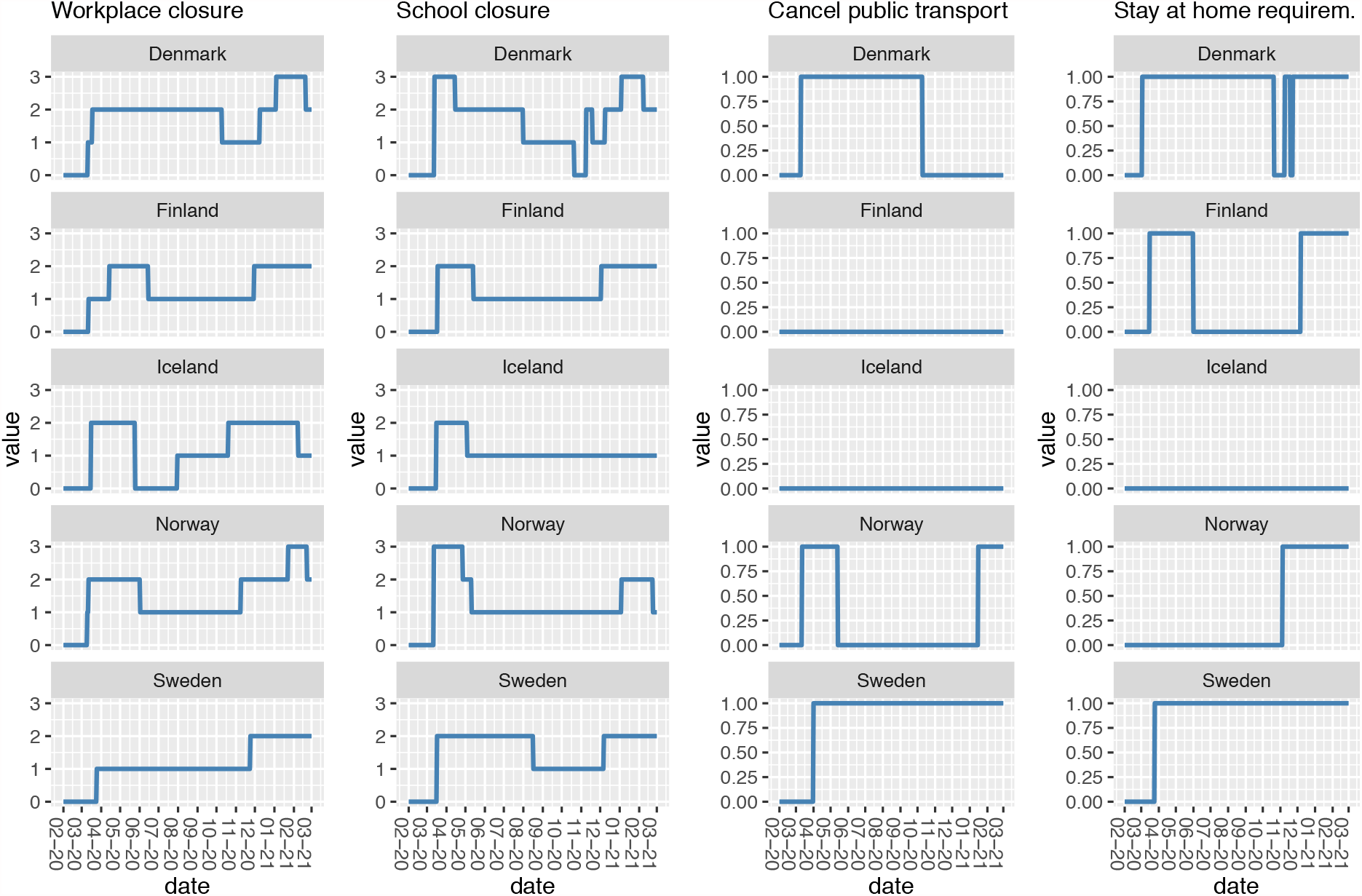
Information on indicators of government response between February 2020 through February 2021 by country regarding (i) workplace closure, (ii) school closure, (iii) cancelation of public transportation, and (iv) stay at home requirements. All indicators are represented using ordinal scales, showing the highest value in the countries at each time point. If local measures are used, these values may apply to a limited geographic area. Workplace closure: 0: no; 1: recommend work from home; 2: require closing for some sectors; 3: require closing for all-but-essential sectors; School closure: 0: no; 1: recommend closing with alteration; 2: require closing some levels; 3: require closing all levels; Cancel public transport: 0: no; 1: recommend closing; 2: require closing; Stay at home requirements: 0: no; 1: recommended not leaving house.

**Supplementary Figure S4.**
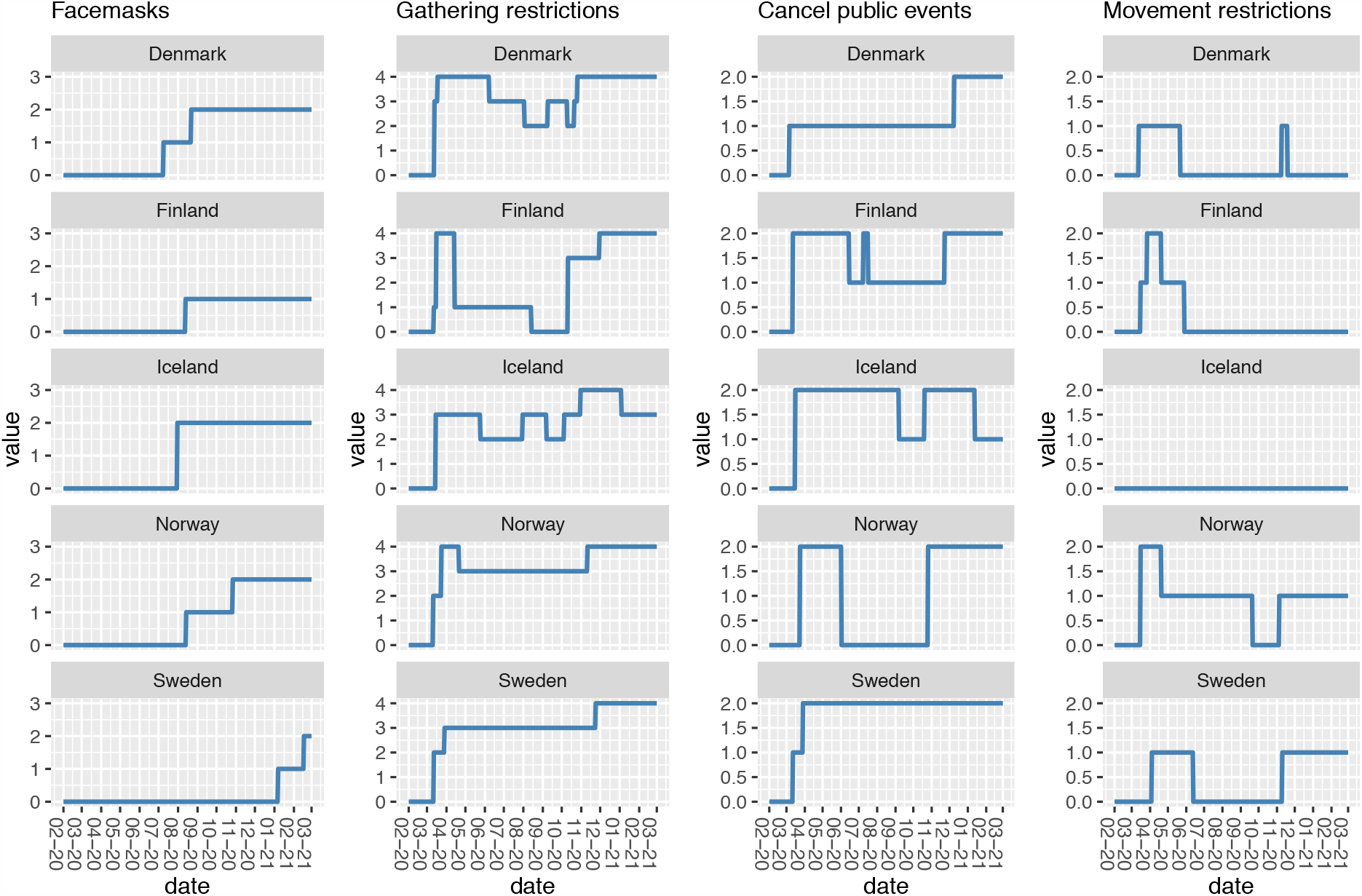
Information on indicators of government response between February 2020 through February 2021 by country regarding (i) facemasks, (ii) gathering restrictions, (iii) cancelation of public events, and (iv) restrictions on movement. All indicators are represented using ordinal scales, showing the highest value in the countries at each time point. If local measures are used, these values may apply to a limited geographic area. Face masks: 0: no; 1: recommend; 2: required in specified public spaces; 3: required in all public spaces when social distancing not possible; Restrictions on gatherings: 0: no; 1: >1000; 2: >100; 3: >10; 4: 10 or less; Cancel public events: 0: no; 1: recommended; 2: required closing; Restrictions on movements: 0: no; 1: recommended no travel between regions; 2: internal movement restrictions.

**Supplementary Figure S5.**
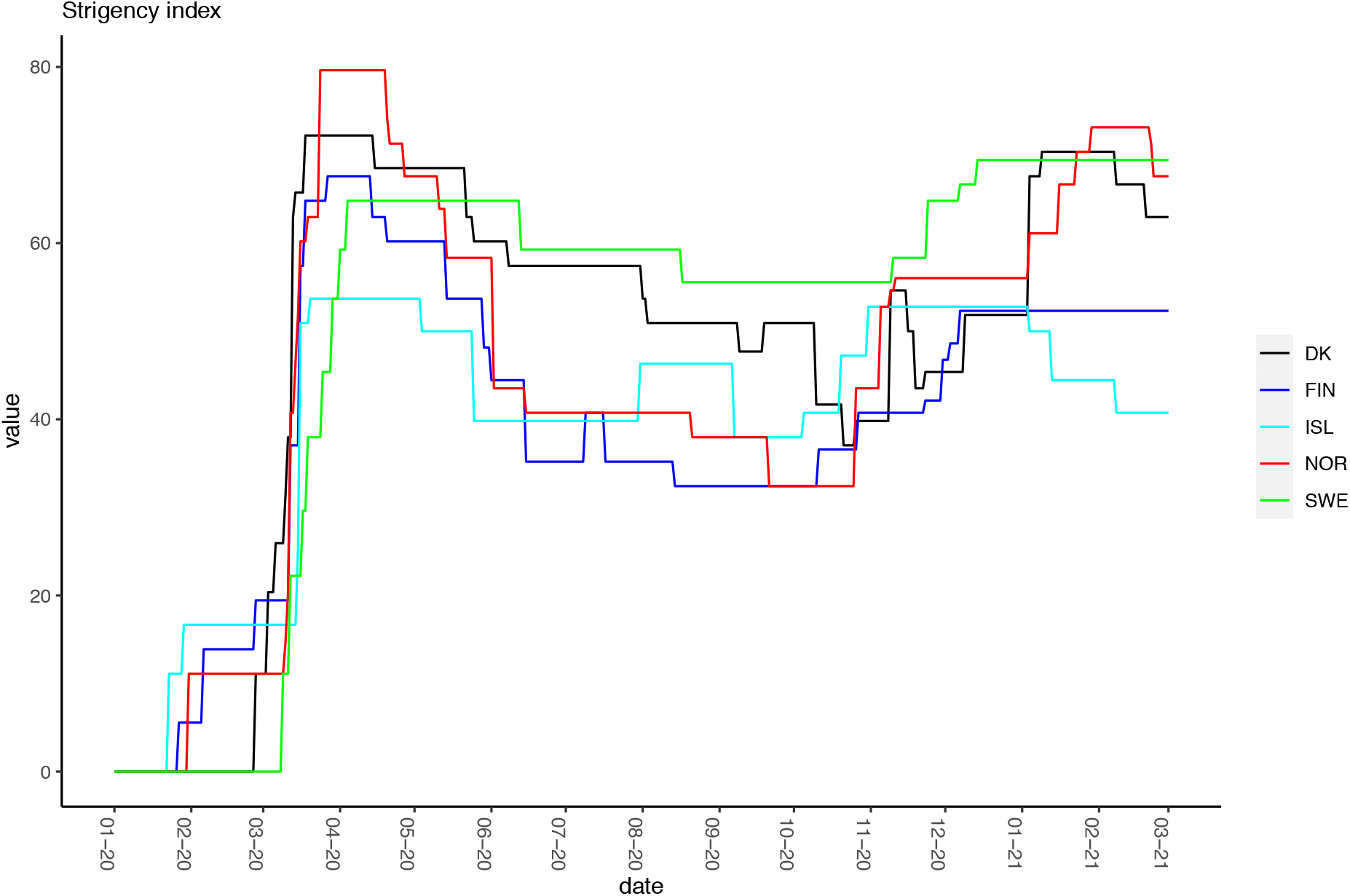
Stringency index of government responses between February 2020 through March 2021 by country. The index is an average of 9 indicators and adjusted for geographic scope. The following indicators are included: school closure, workplace closure, cancel public events; restrictions on gatherings; close public transport, stay at home requirements, restrictions on internal movements, international travel controls, public information campaigns.

**Supplementary Figure S6.**
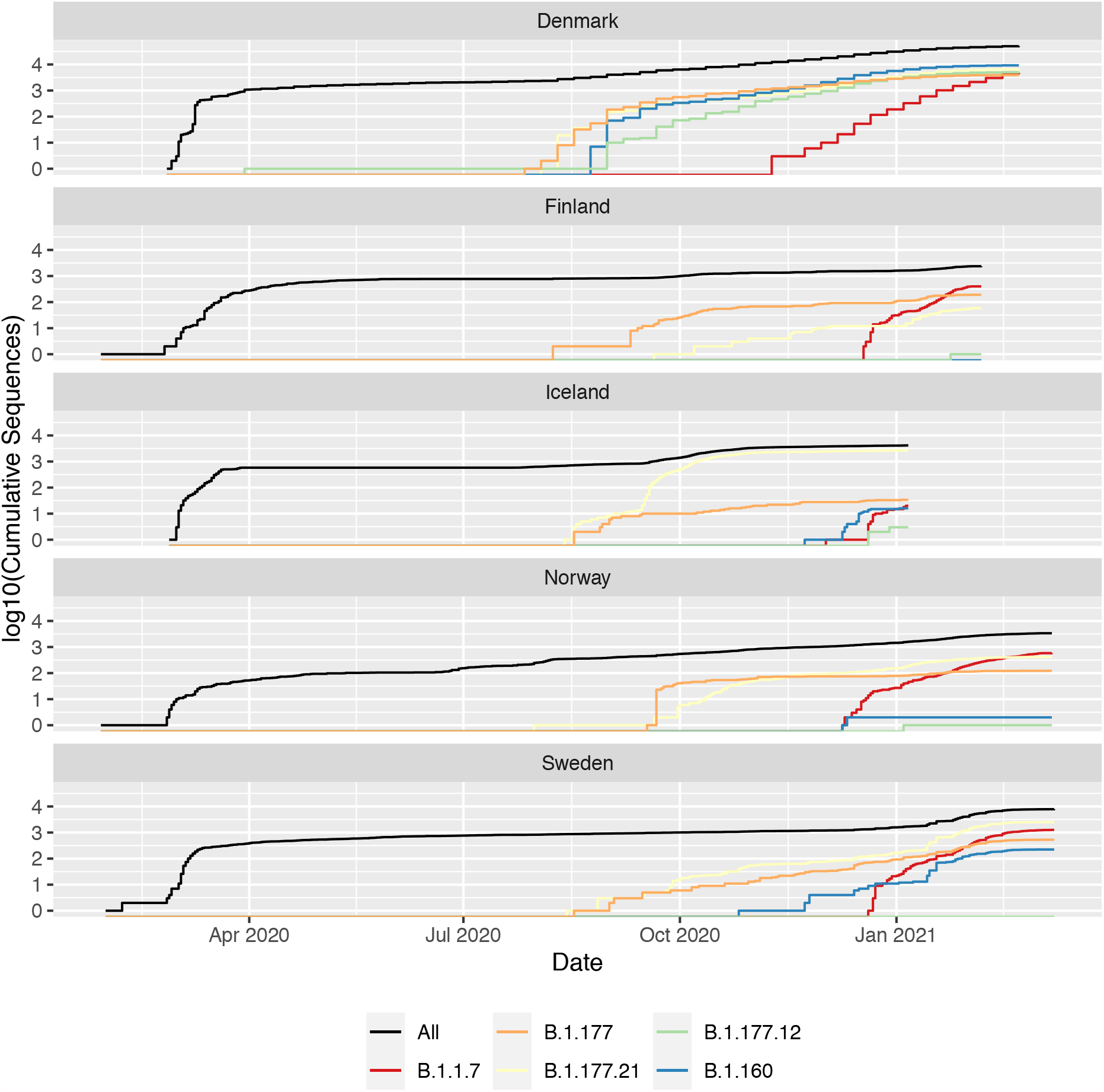
Log number of genomes in each of the five most common pango lineages and B1.1.7 over time in the complete dataset.

**Supplementary Figure S7.**
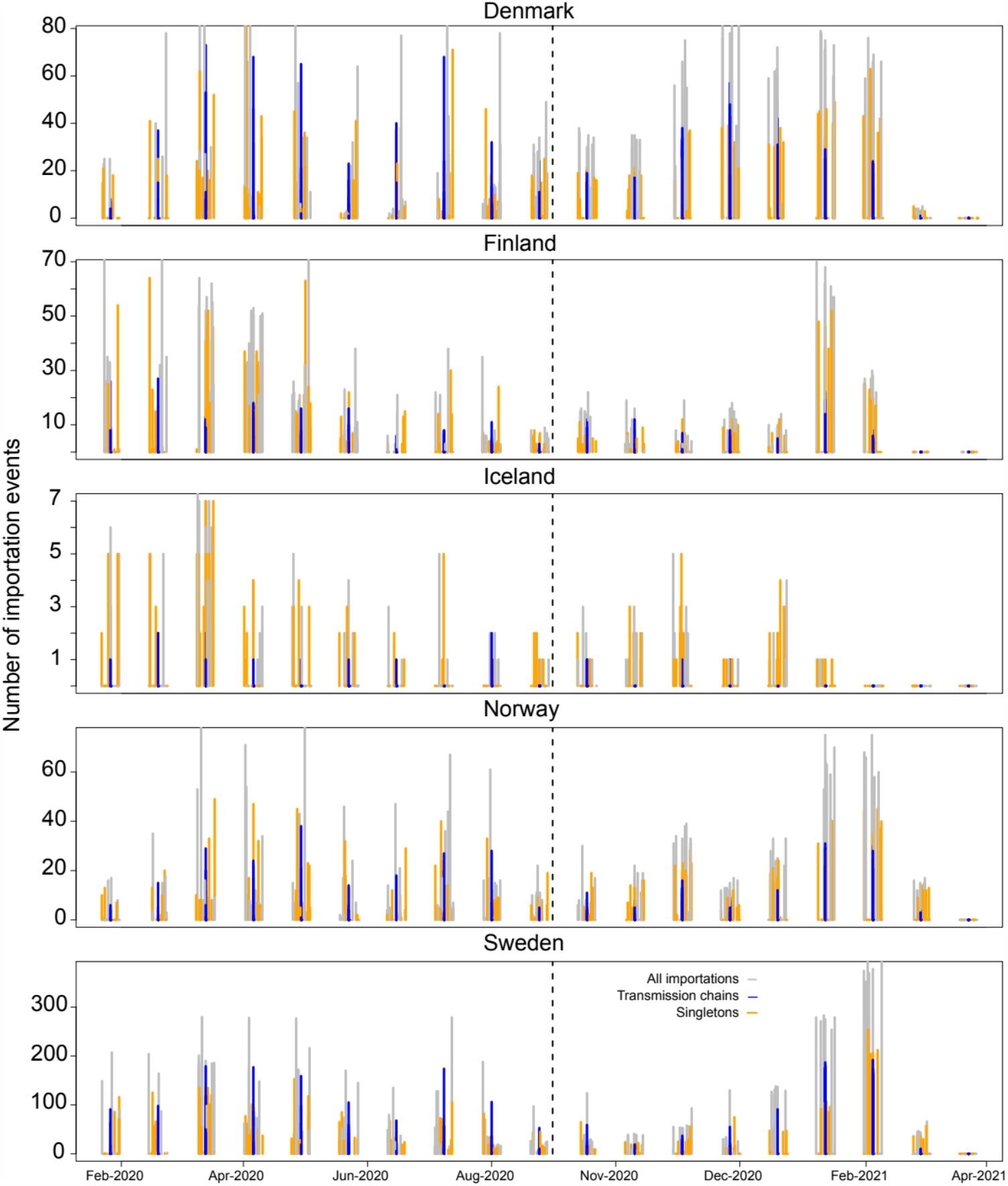
Number of putative introductions identified by detecting transmission chains (monophyletic groups of at least 2 genomes from a Nordic country) or singletons (genomes from a Nordic country sitting outside a transmission chains). Grey bars correspond to the total number of introductions per fortnight. Orange bars are for singletons and those in blue are for transmission chains. Here we included all importation events detected across 10 phylogenetic trees, where each tree contains Nordic genomes sampled relative to prevalence, such that sequencing intensity is equivalent.

**Supplementary Figure S8.**
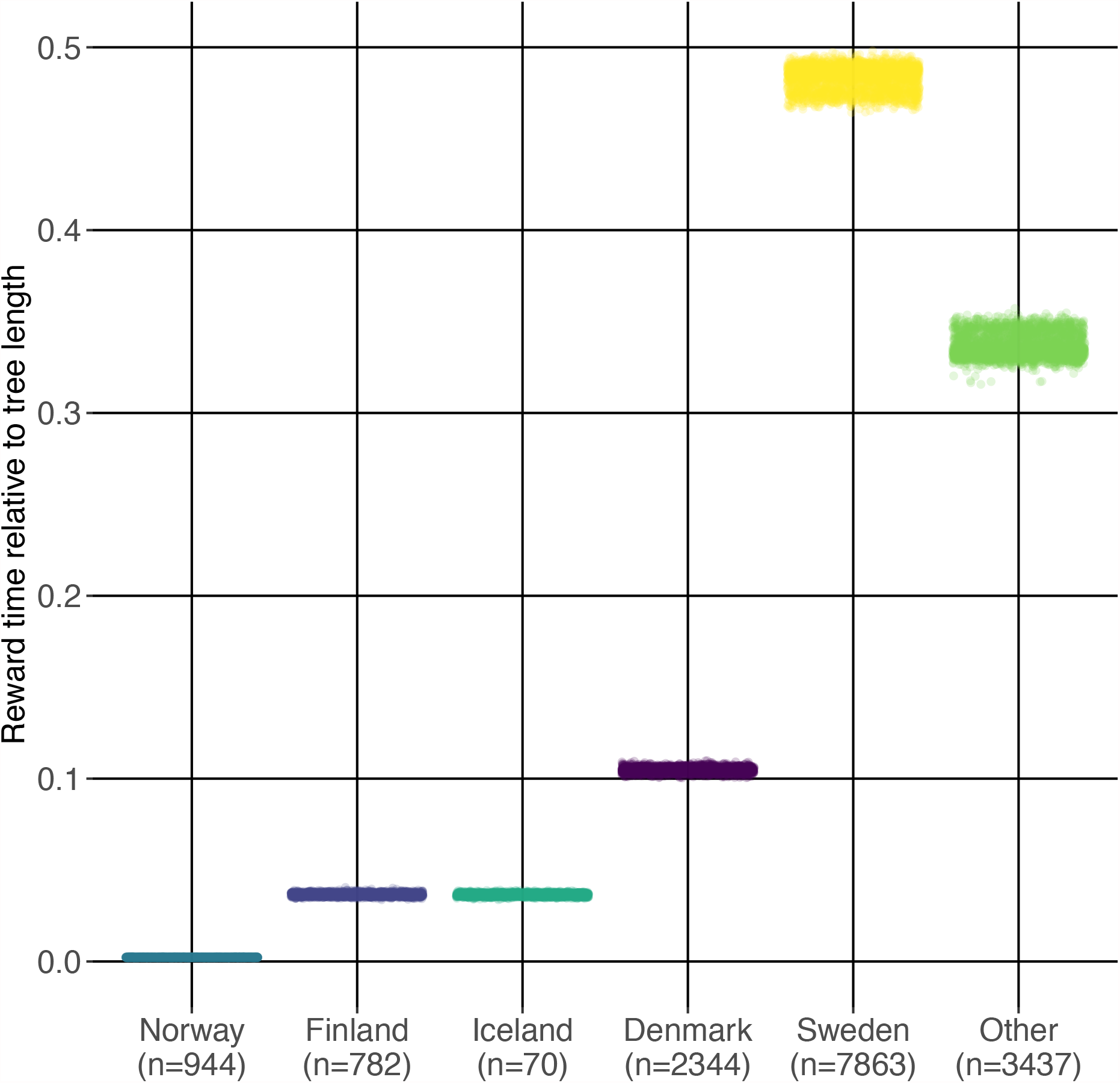
Jittered markov rewards for each Nordic country and for the global diversity in the data (labelled as “other”). The Markov rewards correspond to the portion of time that all lineages spend at either of the six states in the stochastic mapping. The y-axis is the Markov rewards as a proportion of total tree length. Note that countries with most exportation events, Sweden and Denmark are also occupy the largest portion of the tree.

**Supplementary Table S1**. Zip file containing a table including all GIASID accession numbers and replicate information for all sequences SARS-CoV-2 used in the analyses.

**Supplementary Table S2**. Summary table of mitigation responses taken by the Nordic countries until 2021-03-25 according to the European Centre for Disease Control.

